# Comparison of Oscillometry with Lung Function Parameters between Bronchial Asthma with Airflow Obstruction and COPD Patients

**DOI:** 10.1101/2024.10.25.24316127

**Authors:** Sajal De, Aakansha Ashok Sarda

**Author notes:** **Correspondence and reprint requests:** Dr Sajal De, Department of Pulmonary Medicine, All India Institute of Medical Sciences, Raipur, Chhattisgarh, India.

## Abstract

**Background:** Lung oscillometry is an emerging lung function test for assessing obstructive airway disease. Comparisons of oscillometry parameters and their bronchodilator responsiveness (BDR) between bronchial asthma and chronic obstructive pulmonary disease (COPD) patients are limited.

**Research Question:** Do oscillometry parameters and their BDR differ between stable asthma and COPD patients with similar severity of airflow obstruction?

**Study Design and Methods:** We included 467 consecutive adult patients with a clinical history of asthma (n=187) or COPD (n=280). Oscillometry, spirometry, and body plethysmography were performed before and after inhaling 400 μg of salbutamol. Patients were stratified based on the severity of airflow obstruction in spirometry. The z scores of the oscillometry parameters were used for the comparison. The BDR of oscillometry parameters with other lung function parameters was also compared.

**Results:** The average age of the study population was 54.9 years, and 76.4% were male. COPD patients were older, had a greater number of smokers, and had poorer lung function. The magnitude of oscillometry parameters worsened with increasing severity of airflow obstruction, regardless of the underlying disease. Asthma patients, particularly those with moderate and severe airway obstruction, had significantly higher R5 and R19 than COPD patients. The within- and whole-breath X5 of asthma were not different from those of COPD patients with similar severities of airflow obstruction. Expiratory flow limitation at tidal breaths (ΔX5 > 0.28 kPa/L/s) was observed in both asthma and COPD patients across all severities of airflow obstruction. The proportion of BDR in oscillometry was significantly lower than that in spirometry for both asthma (35.3% vs. 57.1%; p<0.01) and COPD patients (19.3% vs. 47.1%; p=0.02).

**Interpretation:** Oscillometry parameters except for R5 and R19 did not differ between asthma and COPD patients with similar severities of airflow obstruction. Similar to spirometry, COPD patients had lower BDR in oscillometry than asthma patients.

**Take-home Points:** *Study Question:* Are oscillometry parameters and their bronchodilator responsiveness different between bronchial asthma and COPD patients with similar severities of airflow obstruction?

*Results:* We compared the FOT between 187 bronchial asthma and 280 COPD patients. Except for R5 and R19, the severity and distribution of high oscillometry parameters did not differ between asthma and COPD patients.

*Interpretation:* The severity of oscillometry abnormalities is primarily determined by the severity of airflow obstruction, not the underlying disease.

## BACKGROUND

Asthma and chronic obstructive pulmonary disease (COPD) are major obstructive airway diseases that affect millions.^1,2^ Both diseases are characterized by chronic inflammation leading to airflow obstruction. However, the types of airway inflammation, pathophysiological changes, and bronchodilator responsiveness (BDR) of spirometry differ.^3^

Oscillometry is an emerging noninvasive technique for assessing small airway dysfunction. It superimposes multiple oscillations on tidal breaths to measure respiratory system resistance (Rrs) and reactance (Xrs). Collapsing the small airways during tidal breathing increases the difference between inspiratory and expiratory Xrs.

Spirometry is the gold standard for diagnosing airflow obstruction and assessing BDR. Forced expiratory volume in 1 s (FEV_1_) and forced vital capacity (FVC) measured by spirometry primarily reflect the functionality of medium to large airways. The correlation between oscillometry parameters and spirometry parameters is generally weak.^4^ The BDR criteria for spirometry defined by the European Respiratory Society (ERS)/American Thoracic Society (ATS) in 2022 differ from their previous criteria published in 1991.^5,6^

Patients with obstructive airway diseases may have high specific airway resistance (sRaw) and residual volume (RV). A reduction in sRaw by ≥50% after inhalation of salbutamol confirms BDR of sRawwith certainty.^7^ A more than 20% decrease in the RV after inhalation of salbutamol had 70% sensitivity and 60% specificity for the BDR of the RV based on FEV_1_.^8^

The primary objective of our study was to compare oscillometry parameters (within- and whole-breath) and their bronchodilator responsiveness with other lung function parameters between stable asthma with airflow obstruction and COPD patients with similar severity of airflow obstruction. The secondary objective was to compare the BDR of the spirometry indices of asthma and COPD patients according to the ERS/ATS criteria published in 1991 and 2022 and their agreement with the BDR of oscillometry parameters.

### Study Design and Methods

#### Population

This retrospective study included clinically diagnosed adult asthma and COPD patients who underwent lung function testing at our department between July 2021 and December 2023. The Institutional Ethics Committee approved the study protocol (No: 4705/IEC-AIIMSRPR/2024). The clinical diagnosis of asthma was based on a history of respiratory symptoms, e.g., wheezing, shortness of breath, chest tightness, and cough, which varied over time and in intensity as per the GINA guidelines.^1^ The clinical diagnosis of COPD was based on the patient’s clinical history and symptoms, including dyspnea on exertion, as per the GOLD guidelines, and postbronchodilator FEV_1_/FVC <70%.^2^ All lung functions were performed before and 15–20 min after inhaling 400 μg of salbutamol by a metered-dose inhaler through a spacer to assess the BDR.

#### Oscillometry

The Resmon Pro Full (V3; RestechSrl, Milan, Italy) was used for oscillometry using 5, 11, and 19 Hz sinusoidal signals. Oscillometry was performed according to the ERS technical standard.^9^ The tests were carried out while the participants were sitting, wearing a nose clip, and with their cheeks supported. At least three tests were performed, each continued until ten accepted breaths were recorded. The means of three trials with coefficients of variation for R5 <10% were used for the analysis. The parameters included were whole-breath Rrs at 5 Hz (R5), within- and whole-breath Xrs at 5 Hz (X5); the difference between inspiratory and expiratory reactance at 5 Hz (ΔX5); and the difference in Rrs between 5 and 19 Hz (R5-19), the resonant frequency (Fres), and the area above the Xrs curve from 5 Hz to the resonant frequency (AX). The severity of oscillometry parameters was compared using the z scores recommended by the ERS technical standards.^9^ The z scores were computed according to the predictive equation of oscillometry parameters for the local population.^10^ R5 and R5-19 were classified as “high” if their z scores were > 1.64. X5 were classified as “high” if their z scores were > -1.64. Severity was classified based on z score, i.e., mild (> 1.64 and ≤2.5), moderate (> 2.5 and ≤ 4), and severe (> 4) impairment of R5 and mild (> - 1.64 and ≤ -2.5) moderate (> - 2.5 and ≤- 4) and severe (>- 4) impairment of X5. Owing to the nonavailability of the z scores of Fres and AX of the local population, we considered Fres > 18 Hz and AX > 30 cmH_2_O/L/s as high. BDR was defined according to the ERS technical standard, i.e., a 40% decrease in R5, a 50% decrease in X5, and an 80% decrease in AX.^9^ The presence of BDR in R5, X5, or AX was defined as the BDR of oscillometry. Tidal expiratory flow limitation (EFL_T_) was defined as ΔX5 > 0.28 kPa/L/s. We defined paradoxical responsiveness (PR) in oscillometry as a 40% increase in R5 or a 50% increase in X5 after bronchodilator use.

A receiver operating characteristic (ROC) analysis was conducted to determine the cutoff for the BDR of oscillometry parameters in terms of changes in the z score and absolute values using the cutoff for BDR as recommended by the ERS technical standards.^9^

### Spirometry and body plethysmography

Spirometry and body plethysmography were performed according to the ERS/ATS recommendations using a PowerCube Body^+^ (GANSHORN Medizin Electronic, Germany) while the patients were sitting and wearing a nose clip.^5^ To assess the BDR of spirometry, we used the ERS/ATS guidelines published in 1991 (i.e., ≥12% and ≥200 mL increase in FEV_1_ and/or FVC from baseline) and in 2022 (i.e., >10% relative to the predicted value for FEV_1_ and/or FVC).^5,6^ The Global Lung Function Initiative equations were used to assess the BDR as per the ERS/ATS-2022 guidelines.^11^ A PR on spirometry was defined as a ≥12% and 200 mL reduction in FEV_1,_ FVC, or both. The predictive equations of spirometry parameters for the Indian population were used to calculate the FEV_1_% predicted.^12^ Without the z-score of spirometry parameters of the local population, we stratified the severity of airflow obstruction based on prebronchodilator FEV_1_% predicted as per the ERS/ATS-1991 guidelines, i.e., FEV_1_% predicted ≥70% mild; <70% to ≥60% moderate; <60% to ≥50% to moderately severe; <50% to ≥35% severe; and <35% very severe. ^5^

A postbronchodilator increase in maximal mid-expiratory flow (MMEF) ≥30% of baseline was used to define BDR in MMEF.^13^ A >20% decrease in RV and a ≥50% reduction in sRaw were defined as BDR in body plethysmography.^7,8^

### Statistical methods

The data were analyzed using IBM SPSS statistics for Windows (Version 23.0, Armonk, NY: IBM crop). Categorical variables are presented as numbers and percentages. Continuous data are presented as the mean ± standard deviation. Differences in proportions were tested using the Pearson χ^2^ test. The student’s t-test was used to compare the variables between the two groups. A p-value of <0.05 was statistically significant. The consistency between the different parameters was evaluated using Cohen’s kappa (κ). The values of κ 0–0.20, 0.21–0.40, 0.41– 0.60, and 0.61–0.80 indicate slight, fair, moderate, and substantial agreement, respectively. Univariate (unadjusted odds ratio) analysis was performed to identify the predictors (age, sex, disease category, severity of airflow obstruction, BDR of FVC, FEV_1_, MMEF, RV, sRaw) for the BDR of oscillometry parameters. Based on a p-value <0.20 in the univariate analysis, a multivariate logistic regression (adjusted odds ratio and confidence intervals) analysis was performed.

## RESULTS

### Study population

Lung function data of 532 clinically diagnosed adult asthma (n=248) and COPD (n=284) patients were extracted from our records. Each spirometry recording was visually inspected for technical errors. Only technically acceptable data were included in the analysis. We excluded 61 patients with clinically diagnosed asthma who did not have airflow obstruction, i.e., prebronchodilator FEV_1_/FVC >75%, from the analysis. Four COPD patients were excluded from the analysis because their postbronchodilator FEV_1_/FVC was >70%. The final study population comprised 467 patients with 187 asthma and 280 COPD patients. The mean (±SD) age of the cohort was 54.9 (15.1) years. Among the 467 patients, 76.4% were male. The anthropometric measurements and spirometry parameters of asthma and COPD patients are compared in Table 1. COPD patients were mostly smokers (79.3%), had a greater male predominance (91.4%), were older, and had poorer lung function (FEV_1_% predicted 49.2% vs. 67.9%, p<0.01).

**Table 1.** The anthropometric and pre-bronchodilator lung function parameters of the cohort.

| Variables | Asthma<br>(n=187) | COPD<br>(n=280) | Total (N=467) |
| --- | --- | --- | --- |
| Male, n (%) | 101 (54) | 256 (91.4)* | 357 (76.4) |
| Age, years | 42.6±13.5 | 63.2±9.4* | 54.9±15.1 |
| Smoking, n (%) | 14 (7.5) | 222 (79.3)* | 236 (50.5) |
| FEV<sub>1</sub>% predicted<sup>#</sup> | 67.9±18.9 | 49.2±18.5* | 56.7±20.8 |
| FVC % predicted<sup>#</sup> | 81.2±18.3 | 65.3±16.6* | 71.7±18.9 |
| FEV<sub>1</sub>/FVC% | 64.7±8.5 | 52.0±9.4* | 57.1±10.9 |
| sRaw, kPa/s | 2.82±2.54 | 4.59±2.9* | 4.04±2.92 |
| RV/TLC, % predicted | 47.8±14.4 | 62±9.7* | 57.6±13.1 |
| High R5 only, n (%) | 27 (14.4) | 12 (4.3)* | 39 (8.4) |
| High X5 only, n (%) | 2 (1.1) | 34 (12.1)* | 36 (7.7) |
| Both R5 and X5 high, n (%) | 91 (48.7) | 194 (69.3)* | 285 (61) |

Notes: <sup>#</sup>: calculated as per the predicted equation for the Indian population; \*: p-value <0.01 Abbreviations: FEV<sub>1</sub>: forced expiratory volume 1 s; FVC: forced vital capacity; sRaw: specific airway resistance; RV/TLC: ratio of residual volume and total lung capacity, High R5: resistance at 5Hz >1.64; High X5: reactance at 5 Hz>-1.64.

### Oscillometry parameters

The oscillometry parameters of asthma and COPD patients were compared according to the severity of airflow obstruction determined by spirometry (Table 2). The majority of asthma patients had mild airflow obstruction (49.7%). With increasing severity of airflow obstruction, the magnitude of oscillometry parameters progressively worsened in both asthma and COPD patients. Asthma patients, particularly those with moderate to severe airflow obstruction, had significantly higher R5 and R19 than COPD patients with similar severity of airflow obstruction. The magnitude of R5-19 did not differ between asthma and COPD patients, regardless of the severity of airflow obstruction. The within- and whole-breath X5 values of asthma were not significantly different from those of COPD patients across all severities of airflow obstruction. The AX of asthma was not different from that of COPD patients across all severities of airflow obstruction. Except for mild airflow obstruction, the Fres of asthma was also not different from that of COPD patients. The prevalence of EFL_T_ between asthma and COPD patients was not different, except for very severe airflow obstruction.

**Table 2.** Comparison of pre-bronchodilator oscillometry parameters between bronchial asthma and COPD patients stratified by severity of airflow obstruction.

|  | Mild airflow obstruction<br>(n=128) |  |  | Moderate airflow obstruction<br>(n=73) |  |  | Moderately severe airflow obstruction<br>(n=70) |  |  | Severe airflow obstruction<br>(n=117) |  |  | Very severe airflow obstruction<br>(n=79) |  |  |
| --- | --- | --- | --- | --- | --- | --- | --- | --- | --- | --- | --- | --- | --- | --- | --- |
| Variables | Asthma<br>(n=93) | COPD<br>(n=35) | p-value | Asthma<br>(n=33) | COPD<br>(n=40) | p-value | Asthma<br>(n=28) | COPD<br>(n=42) | p-value | Asthma<br>(n=22) | COPD<br>(n=95) | p-value | Asthma<br>(n=11) | COPD<br>(n=68) | p-value |
| Male, n (%) | 52<br>(55.9) | 29*<br>(82.9) | <0.01 | 20<br>(60.6) | 39*<br>(97.5) | <0.01 | 14 (50) | 39*<br>(92.9) | <0.01 | 8<br>(36.4) | 83*<br>(87.4) | <0.001 | 7<br>(63.6) | 66*<br>(97.1) | <0.01 |
| R5 total, z-score | 2.02±2.26 | 1.86±2.14 | 0.71 | 2.83±2.12 | 2.16±2.29 | 0.19 | 4.49±2.31 | 2.86±1.97* | <0.01 | 5.52±3.5 | 3.71±2.17 | <0.01 | 5.41±2.13 | 4.11±2.09 | 0.06 |
| High R5, n (%) | 44<br>(47.3) | 19<br>(54.3) | 0.30 | 21<br>(63.6) | 20<br>(50) | 0.18 | 24<br>(85.7) | 28<br>(66.7) | 0.06 | 18<br>(81.8) | 78<br>(82.1) | 0.59 | 11<br>(100) | 61<br>(89.7) | 0.33 |
| R19 total, z-score | 2.07±1.93 | 1.77±2.06 | 0.44 | 2.40±1.72 | 1.58±1.90 | 0.06 | 3.22±1.7 | 1.98±1.71* | <0.01 | 3.79±2.53 | 2.37±1.87* | <0.01 | 3.59±2.01 | 2.53±1.71 | 0.07 |
| High R19, n (%) | 44<br>(47.3) | 19<br>(54.3) | 0.31 | 21<br>(63.6) | 20<br>(50) | 0.18 | 24<br>(85.7) | 28<br>(66.7) | 0.06 | 18<br>(81.8) | 78<br>(82.1) | 0.59 | 11<br>(100) | 61<br>(89.7) | 0.33 |
| R5-19, z-score | 0.81±1.85 | 1.07±2.24 | 0.49 | 1.92±1.95 | 2.18±2.33 | 0.62 | 3.51±2.56 | 2.99±2.2 | 0.37 | 3.41±2.43 | 3.97±2.21 | 0.29 | 5.37±2.91 | 5.35±2.75 | 0.99 |
| High R5-19, n (%) | 22<br>(23.7) | 9<br>(25.7) | 0.49 | 15<br>(45.5) | 24<br>(60) | 0.16 | 21<br>(75) | 29<br>(69) | 0.39 | 17<br>(77.3) | 81<br>(85.3) | 0.27 | 10<br>(90.9) | 64<br>(94.1) | 0.54 |
| X5insp, z-score | -0.76±1.19 | -0.94±1.01 | 0.43 | -1.37±1.25 | -1.63±1.35 | 0.39 | -2.53±1.57 | -2.28±1.57 | 0.50 | -2.35±1.59 | -3.02±1.73 | 0.09 | -<br>4.02±2.22 | -4.11±2.22 | 0.89 |
| X5exp, z-score | -1.90±3.84 | -2.22±2.63 | 0.66 | -3.38±4.08 | -3.94±4.59 | 0.59 | -6.32±4.67 | -6.68±6 | 0.79 | -9.03±7.48 | -8.80±5.08 | 0.86 | -<br>9.68±5.71 | -<br>10.23±5.26 | 0.75 |
| X5total, z-score | -1.32±2.73 | -1.69±2.11 | 0.47 | -2.53±2.96 | -3.26±3.84 | 0.37 | -4.81±3.45 | -5.29±4.68 | 0.64 | -6.38±5.11 | -7.05±3.98 | 0.5 | -<br>7.75±4.64 | -8.88±4.55 | 0.45 |
| High X5, n (%) | 25<br>(26.9%) | 16*<br>(45.7) | 0.04 | 18<br>(54.5) | 26<br>(65) | 0.25 | 21<br>(75) | 33<br>(78.6) | 0.47 | 18<br>(81.8) | 88<br>(92.6) | 0.13 | 11<br>(100) | 65<br>(95.6) | 0.64 |
| ΔX5 (cm H<sub>2</sub>O/L/s) | 0.61±1.74 | 0.85±1.36 | 0.47 | 1.22±1.95 | 1.58±2.38 | 0.48 | 2.15±2.12 | 2.89±3.23 | 0.29 | 3.82±3.81 | 3.68±2.46 | 0.83 | 3.26±2.49 | 3.99±2.76 | 0.41 |
| EFL<sub>T</sub>, n (%) | 9<br>(9.7) | 2<br>(5.7) | 0.38 | 5<br>(15.2) | 7<br>(17.5) | 0.52 | 10<br>(35.7) | 15<br>(35.7) | 0.59 | 11<br>(50.0) | 56<br>(58.9) | 0.29 | 3<br>(27.3) | 41*<br>(60.3) | 0.04 |
| AX (cm H<sub>2</sub>O/L/s) | 16.67±20.33 | 20.59±16 | 0.31 | 25.98±21.36 | 30.1±22.27 | 0.43 | 42.51±25.46 | 42.53±25.37 | 0.99 | 56.24±37.02 | 52.4±24.46 | 0.55 | 62.19±27.67 | 60.64±24.79 | 0.85 |
| High Ax, n (%) | 14<br>(15.1) | 8<br>(22.9) | 0.22 | 10<br>(30.3) | 18<br>(45) | 0.15 | 18<br>(64.3) | 27<br>(64.3) | 0.60 | 17<br>(77.3) | 75<br>(78.9) | 0.53 | 10<br>(90.9) | 61<br>(89.7) | 0.69 |
| Fres, Hz | 18.97±6.45 | 22.24±6.4 | 0.01 | 22.7±6.96 | 25.14±6.28 | 0.12 | 25.81±5.5 | 27.03±5.57 | 0.37 | 26.27±6.44 | 27.44±4.3 | 0.30 | 28.88±3. | 28.13±3.9 | 0.55 |
|  |  | 9* |  |  |  |  | 3 |  |  |  | 2 |  | 29 | 2 |  |
| High Fres,<br>n (%) | 53<br>(57) | 25<br>(71.4) | 0.09 | 25<br>(75.8) | 33<br>(82.5) | 0.34 | 26<br>(92.9) | 38<br>(90.5) | 0.54 | 19<br>(86.4) | 93<br>(97.9) | 0.05 | 11<br>(100) | 68<br>(100) | - |

Notes: \*: p-value<0.05
Abbreviations: R5: whole-breath respiratory system resistance at 5 Hz; R19: whole-breath respiratory system resistance at 19 Hz; R5-19: difference in whole-breath resistance at 5 and 19 Hz; X5insp: inspiratory respiratory system reactance at 5 Hz; X5exp: expiratory respiratory system reactance at 5 Hz; X5: whole-breath respiratory system reactance at 5 Hz; ΔX5: difference between inspiratory and expiratory reactance at 5 Hz; AX: reactance area; Fres: resonant frequency; EFL<sub>T</sub>: expiratory flow limitation at tidal breath.

We compared the distribution of R5 and X5 impairment severity between asthma and COPD patients according to the severity of airflow obstruction (Table 3). The distribution of severity of abnormalities in oscillometry parameters was not different between asthma and COPD patients, except for moderately severe airflow obstruction. The prevalence of impairment severity in R5 and X5 increased with increasing severity of airflow obstruction (p<0.001), irrespective of the underlying disease. The ΔX5, i.e., expiratory X5, increased progressively with the severity of airflow obstruction, and it was not different between asthma and COPD patients. We also found that the magnitude and proportion of oscillometry-defined small airway dysfunction (i.e., abnormalities in X5 and R5-19) in asthma were not different from those in COPD patients with similar airflow obstruction.

**Table 3.** Comparison of severity of abnormalities in oscillometry parameters between bronchial asthma and COPD patients across different severity of airflow obstruction.

|  | Mild airflow obstruction<br>(n=128) |  |  | Moderate airflow obstruction<br>(n=73) |  |  | Moderately severe airflow Obstruction<br>(n=70) |  |  | Severe airflow Obstruction<br>(n=117) |  |  | Very severe airflow Obstruction<br>(n=79) |  |  |
| --- | --- | --- | --- | --- | --- | --- | --- | --- | --- | --- | --- | --- | --- | --- | --- |
| Variables | Asthma<br>(n=93) | COPD<br>(n=35) | p-value | Asthma<br>(n=33) | COPD<br>(n=40) | p-value | Asthma<br>(n=28) | COPD<br>(n=42) | p-value | Asthma<br>(n=22) | COPD<br>(n=95) | p-value | Asthma<br>(n=11) | COPD<br>(n=68) | p-value |
| **Severity of R5** |  |  | 0.06 |  |  | 0.39 |  |  | 0.03 |  |  | 0.24 |  |  | 0.25 |
| Mild, n (%) | 16 (17.2) | 5 (14.3) |  | 5 (15.2) | 5 (12.5) |  | 1 (3.6) | 8 (19) |  | 1 (4.5) | 12 (12.6) |  | 0 | 6 (8.8) |  |
| Moderate, n (%) | 11 (11.8) | 11 (31.4) |  | 7 (21.2) | 10 (25) |  | 7 (25) | 8 (19) |  | 3 (13.6) | 26 (27.4) |  | 2 (18.2) | 20 (29.4) |  |
| Severe, n (%) | 17 (18.3) | 3 (8.6) |  | 9 (27.3) | 5 (12.5) |  | 16 (57.1) | 12 (28.6) |  | 14 (63.6) | 40 (42.1) |  | 9 (81.8) | 35 (51.5) |  |
| **Severity of X5** |  |  | 0.06 |  |  | 0.62 |  |  | 0.66 |  |  | 0.44 |  |  | 0.69 |
| Mild, n (%) | 9 (9.7) | 9 (25.7) |  | 5 (15.2) | 5 (12.5) |  | 1 (3.6) | 5 (11.9) |  | 1 (4.5) | 3 (3.2) |  | 1 (9.1) | 2 (2.9) |  |
| Moderate, n (%) | 6 (6.5) | 1 (2.9) |  | 6 (18.2) | 7 (17.5) |  | 5 (17.9) | 6 (14.3) |  | 2 (9.1) | 12 (12.6) |  | 1 (9.1) | 7 (10.3) |  |
| Severe, n (%) | 10 (10.8) | 6 (17.1) |  | 7 (21.2) | 14 (35) |  | 15 (53.6) | 22 (52.4) |  | 15 (68.2) | 73 (76.8) |  | 9 (81.8) | 56 (82.4) |  |

Abbreviations: R5: whole-breath respiratory system resistance at 5 Hz; X5: whole-breath respiratory system reactance at 5 Hz.

### Bronchodilator responsiveness

The BDR of the spirometry, body plethysmography, and oscillometry of asthma and COPD patients are compared in Table 4. Compared with the 1991 criteria, the ERS/ATS-2022 criteria significantly underestimate the BDR of FEV_1_ (43.5% vs. 52.2%, p<0.01), FVC (23.4% vs. 29.9%, p<0.01), and spirometry (47.3% vs. 57.1%, p<0.01) in asthma patients. Adopting the ERS/ATS-2022 criteria also significantly underestimated the BDR of FEV_1_ (16.1% vs. 27.1%, p<0.01), FVC (27.9% vs. 41.4%, p<0.01), and spirometry (31.4% vs. 47.1%, p<0.01) in COPD patients. Regardless of the criteria, the FEV_1_ of asthma patients was significantly greater than that of COPD patients. According to the ERS/ATS-1991 criteria, the FVC of COPD patients was significantly greater than that of asthma patients (41.4% vs. 27.9%, p<0.01), but not according to the 2022 criteria (29.9% vs. 23.4%, p=0.14). The prevalence of BDR, according to the ERS/ATS-1991 criteria, increased with the progression of airflow obstruction in COPD (p<0.01) but not in asthma patients (p=0.51). Agreement on BDR in spirometry between the ERS/ATS-2022 and 1991 criteria for asthma (κ=0.79) and COPD (κ=0.68) was moderate. The sRaw of asthma patients had a significantly greater BDR than that of COPD patients (53.8% vs. 29.7%; p<0.001). The proportion of BDR of sRaw and spirometry according to the ERS/ATS-1991 criteria was not significantly different in asthma patients (53.8% vs. 57.1%), but the agreement was weak (κ =0.44). The BDR of the RV for both asthma (11.1%) and COPD patients (9.4%) was less and was not different (p=0.36).

**Table 4.** Comparison of bronchodilator responsiveness of lung function and oscillometry parameters between bronchial asthma and COPD.

| Variables | Asthma<br>(n=184)<sup>§</sup> | COPD<br>(n=280) |
| --- | --- | --- |
| BDR of FEV<sub>1</sub> (ATS/ERS-1991 criteria)<sup>^</sup>, n (%) | 96 (52.2) | 76 (27.1)* |
| BDR of FEV<sub>1</sub> (ATS/ERS-2022 criteria)<sup>@</sup>, n (%) | 80 (43.5) | 45 (16.1)* |
| BDR of FVC (ATS/ERS-1991 criteria)<sup>^</sup>, n (%) | 55 (29.9) | 116 (41.4)* |
| BDR of FVC (ATS/ERS-2022 criteria)<sup>@</sup>, n (%) | 43 (23.4) | 78 (27.9) |
| BDR of spirometry (ATS/ERS-1991 criteria)<sup>^</sup>, n (%) | 105 (57.1) | 132 (47.1)* |
| BDR of spirometry (ATS/ERS-2022 criteria)<sup>@</sup>, n (%) | 87 (47.3) | 88 (31.4)* |
| BDR of MMEF, n (%) | 106 (57.6) | 62 (22.2) |
| BDR of sRaw, n (%)<sup>#</sup> | 63 (53.8) | 79 (29.7)* |
| BDR of RV, n (%)<sup>#</sup> | 13 (11.1) | 25 (9.4) |
| BDR of R5, n (%) | 52 (27.8) | 32 (11.4)* |
| BDR of X5, n (%) | 52 (27.8) | 46 (16.4)* |
| BDR of AX, n (%) | 34 (18.2) | 11 (3.9)* |
| BDR of Zrs, n (%) | 65 (35.3) | 54 (19.3)* |

Notes: <sup>§</sup>: Bronchodilator responsiveness of three bronchial asthma patients was not performed; <sup>^</sup>: $\geq 12\%$ and $\geq 200$ mL increase; <sup>@</sup>: $>10\%$ relative to the predicted value; <sup>#</sup>: Body plethysmography was carried out in 117 bronchial asthma and 266 COPD patients; \*: p-value $<0.05$ .
Abbreviations: BDR: bronchodilator responsiveness; MMEF: maximal mid-expiratory flow rate; sRaw: specific airway resistance; RV: residual volume; R5: whole-breath respiratory system resistance at 5 Hz; X5: whole-breath respiratory system reactance at 5 Hz; AX: reactance area; Zrs: oscillometry parameters.

The BDR of oscillometry parameters of asthma and COPD patients were significantly lower than those of spirometry, regardless of the criteria used to define BDR by spirometry. Among all oscillometry parameters, AX had the lowest BDR in both asthma (18.2%) and COPD patients (3.9%). The prevalence of BDR of R5 and X5 in asthma was similar, but their concordance was moderate (κ=0.55). The X5 of COPD patients had a significantly higher BDR than the R5 (16.4% vs. 11.4%, p<0.01). The distribution of the BDR of oscillometry parameters across severities of airflow obstruction was not different for either asthma (p=0.62) or COPD (p=0.9) patients. The ERS/ATS-1991 criteria of BDR in spirometry and BDR of FOT had a fair agreement for asthma (κ =0.36) but slight (κ =0.11) for COPD patients.

The PR of spirometry was 2.4%, and it was equally distributed between asthma and COPD patients. PR of R5 was observed in only two patients (0.4%); both were COPD patients. PR of X5 was observed in eight patients (1.7%), and the majority had COPD (n=7).

Multivariate analysis revealed that female sex (adjusted OR, 2.14; 95% CI, 1.02--4.47; p=0.04), the presence of BDR in FEV_1_ (adjusted OR, 2.25; 95% CI, 1.04--4.88; p=0.04), the BDR of MMEF (adjusted OR, 2.85; 95% CI, 1.49--5.48; p=0.002), and the BDR of sRaw (adjusted OR, 6.26; 95% CI, 3.49--11.24; p<0.001) were independently associated with the BDR of oscillometry (Table 5). The severity of airflow obstruction, underlying diseases, and the BDR of FVC were not independently related to the BDR of oscillometry.

**Table 5.**
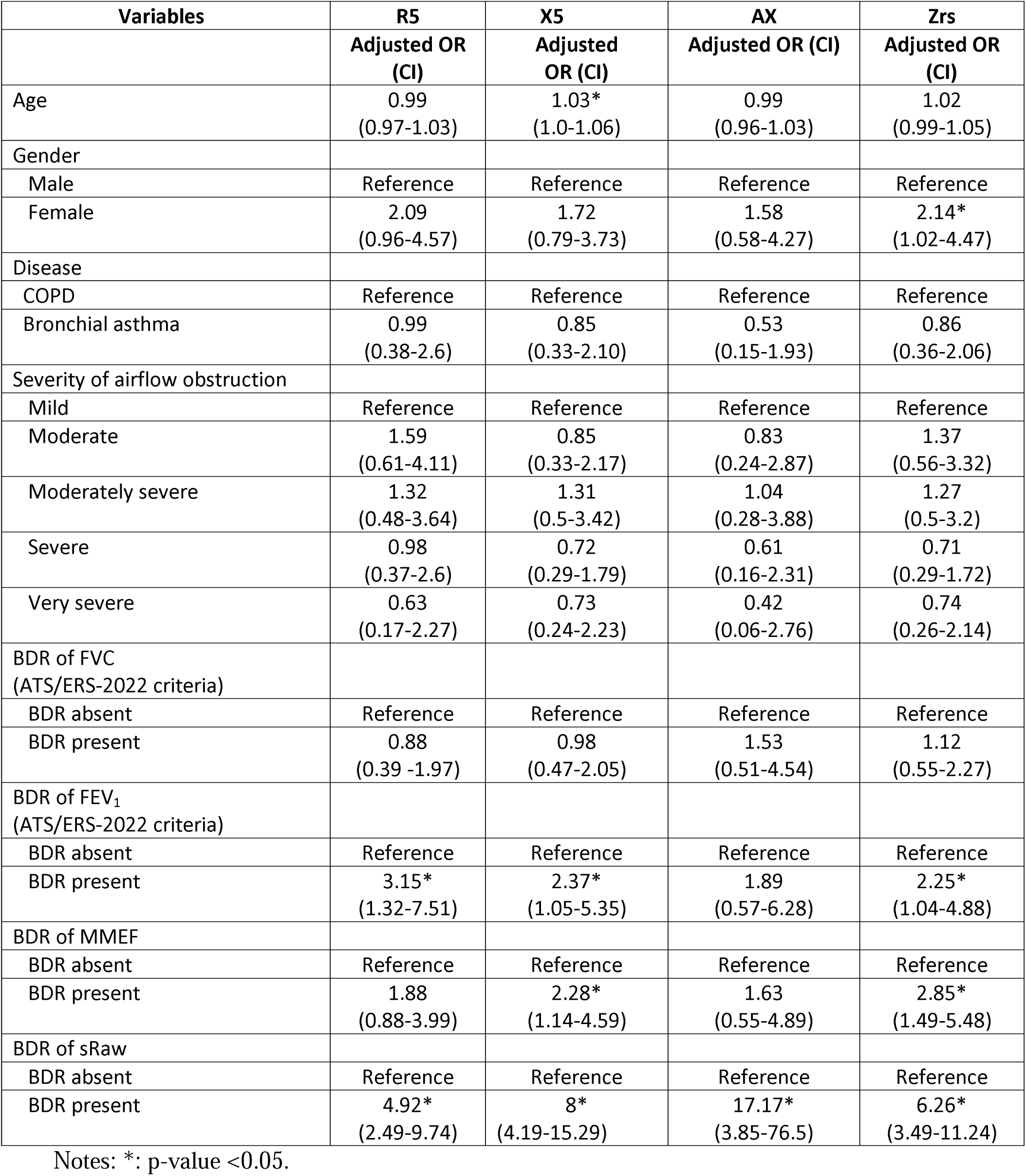
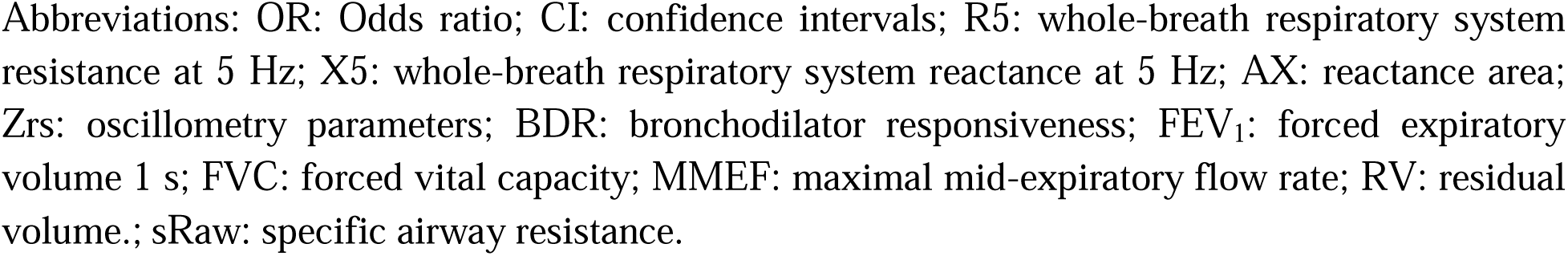
Multivariate analysis for the predictors of bronchodilator responsiveness of oscillometry parameters.

| Variables | R5 | X5 | AX | Zrs |
| --- | --- | --- | --- | --- |
|  | Adjusted OR (CI) | Adjusted OR (CI) | Adjusted OR (CI) | Adjusted OR (CI) |
| Age | 0.99<br>(0.97-1.03) | 1.03*<br>(1.0-1.06) | 0.99<br>(0.96-1.03) | 1.02<br>(0.99-1.05) |
| Gender |  |  |  |  |
| Male | Reference | Reference | Reference | Reference |
| Female | 2.09<br>(0.96-4.57) | 1.72<br>(0.79-3.73) | 1.58<br>(0.58-4.27) | 2.14*<br>(1.02-4.47) |
| Disease |  |  |  |  |
| COPD | Reference | Reference | Reference | Reference |
| Bronchial asthma | 0.99<br>(0.38-2.6) | 0.85<br>(0.33-2.10) | 0.53<br>(0.15-1.93) | 0.86<br>(0.36-2.06) |
| Severity of airflow obstruction |  |  |  |  |
| Mild | Reference | Reference | Reference | Reference |
| Moderate | 1.59<br>(0.61-4.11) | 0.85<br>(0.33-2.17) | 0.83<br>(0.24-2.87) | 1.37<br>(0.56-3.32) |
| Moderately severe | 1.32<br>(0.48-3.64) | 1.31<br>(0.5-3.42) | 1.04<br>(0.28-3.88) | 1.27<br>(0.5-3.2) |
| Severe | 0.98<br>(0.37-2.6) | 0.72<br>(0.29-1.79) | 0.61<br>(0.16-2.31) | 0.71<br>(0.29-1.72) |
| Very severe | 0.63<br>(0.17-2.27) | 0.73<br>(0.24-2.23) | 0.42<br>(0.06-2.76) | 0.74<br>(0.26-2.14) |
| BDR of FVC<br>(ATS/ERS-2022 criteria) |  |  |  |  |
| BDR absent | Reference | Reference | Reference | Reference |
| BDR present | 0.88<br>(0.39 -1.97) | 0.98<br>(0.47-2.05) | 1.53<br>(0.51-4.54) | 1.12<br>(0.55-2.27) |
| BDR of FEV<sub>1</sub><br>(ATS/ERS-2022 criteria) |  |  |  |  |
| BDR absent | Reference | Reference | Reference | Reference |
| BDR present | 3.15*<br>(1.32-7.51) | 2.37*<br>(1.05-5.35) | 1.89<br>(0.57-6.28) | 2.25*<br>(1.04-4.88) |
| BDR of MMEF |  |  |  |  |
| BDR absent | Reference | Reference | Reference | Reference |
| BDR present | 1.88<br>(0.88-3.99) | 2.28*<br>(1.14-4.59) | 1.63<br>(0.55-4.89) | 2.85*<br>(1.49-5.48) |
| BDR of sRaw |  |  |  |  |
| BDR absent | Reference | Reference | Reference | Reference |
| BDR present | 4.92*<br>(2.49-9.74) | 8*<br>(4.19-15.29) | 17.17*<br>(3.85-76.5) | 6.26*<br>(3.49-11.24) |

Notes: \*: p-value <0.05.
Abbreviations: OR: Odds ratio; CI: confidence intervals; R5: whole-breath respiratory system resistance at 5 Hz; X5: whole-breath respiratory system reactance at 5 Hz; AX: reactance area; Zrs: oscillometry parameters; BDR: bronchodilator responsiveness; FEV<sub>1</sub>: forced expiratory volume 1 s; FVC: forced vital capacity; MMEF: maximal mid-expiratory flow rate; RV: residual volume.; sRaw: specific airway resistance.

The decrease of 1.75 z score of R5 had a sensitivity of 75.5% and a specificity of 98.8%, with an AUC of 0.96 for diagnosing BDR using the criterion of a > 40% decrease in R5 (Figure 1). The absolute reduction of 1.68 cmH_2_O/L/s of R5 had an AUC of 0.96, a sensitivity of 75.2%, and a specificity of 99.7% for diagnosing BDR. The reduction in R5-19 of 50% had an AUC of 0.87, with a sensitivity of 64.8% and specificity of 98.4% for diagnosing BDR, using the criteria of >40% reduction in R5. The change in the z score of X5 had an AUC of 0.9, a sensitivity of 96.9%, and a specificity of 65.0% for diagnosing BDR when the criterion of a > 50% decrease in X5. The absolute reduction in X5 of 0.76 cmH_2_O/L/s for X5 had an AUC of 0.91, a sensitivity of 62.6%, and a specificity of 99.6% for diagnosing BDR using the same criterion.

**Figure 1:**
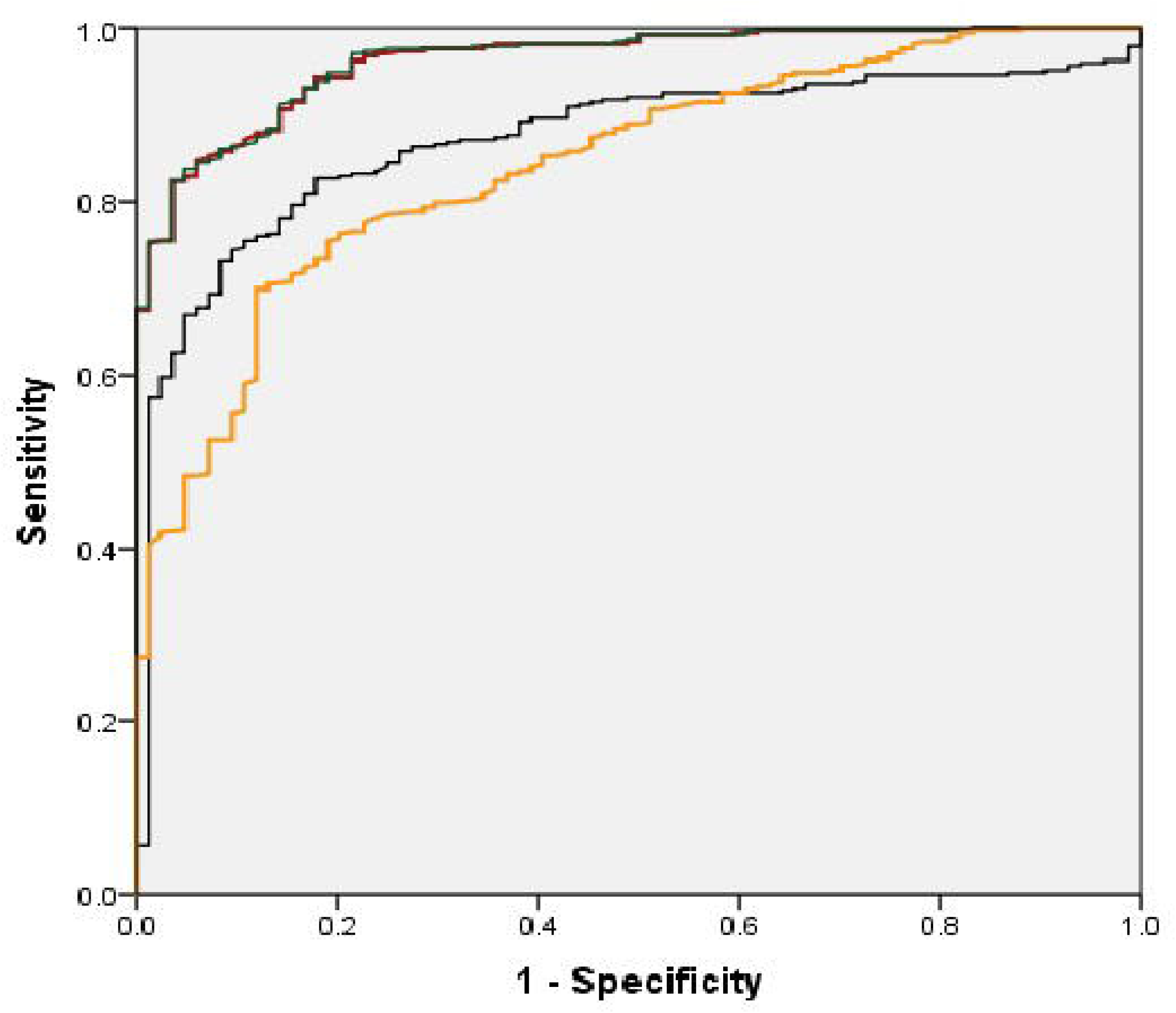
Receiver operating characteristic (ROC) curves of changes in resistance at 5 Hz (R5) and resistance between 5 and 19 Hz (R5-19) are used to identify bronchodilator responsiveness. Red solid line: absolute change in R5; green solid line: change in the z score of R5; black solid line: percentage change in R5--20; yellow solid line: change in R5--19.

## Discussion

We compared the oscillometry parameters and their BDR with other lung function parameters between stable asthma with airflow obstruction and COPD patients with similar severity of airflow obstruction. The magnitude of R5 and R19 was significantly higher in asthma patients with moderate to severe airflow obstruction. The other oscillometry did not differ between asthma and COPD patients, regardless of the severity of airflow obstruction. Adopting the ERS/ATS-2022 criteria significantly reduced the BDR in spirometry of both asthma and COPD patients.

Over the last decade, lung oscillometry has become increasingly popular for evaluating obstructive airway diseases. Similar to our observations, previous studies reported significantly higher R5 and R20 in asthma than in COPD patients.^3,14,-16^ Qi et al. proposed that R5 and R20 are valuable in distinguishing asthma from COPD patients with similar severity of airflow obstructions.^15^ Kanda et al. reported the progressive worsening of oscillometry with increasing severity of airflow obstruction, and only the R20 was significantly higher in asthma than in COPD patients.^16^ Studies have shown that the R5-20 of asthma patients is substantially greater than that of COPD patients, whereas others studies have reported the contradictory.^14–16^ The higher Rrs in asthma is probably because the disease predominantly involves the airways. Kanda et al. reported that the Fres of asthma was lower than that of COPD patients.^15^ Paredi et al. reported that the ΔX5 of COPD was greater than that of asthma patients with similar airflow obstructions, although the difference was insignificant.^17^ They included fewer patients and patients with very severe airflow obstruction were not included. Kamada et al. also reported that ΔX5 of COPD patients was significantly higher than asthma patients.^18^ The unequal distribution of determinants of oscillometry parameters (i.e., age, sex, and standing height) across the groups may impact the comparison. Therefore, we used the z scores of oscillometry parameters for the comparison.

Dellacà et al. suggested that the presence of EFL_T_ is a characteristic feature of severe COPD.^19^ Both asthma and COPD patients in our study had EFL_T_, regardless of the severity of airflow obstruction. We also observed that the presence of EFL_T_ was not exclusive to COPD patients, as suggested by Dellacà et al.^19^

A large population-based European study reported that the BDR in spirometry of asthma and COPD patients according to the ERS/ATS-1991 criteria was lower than ours. This is probably because the lung functions in our study were primarily performed for diagnostic purposes. Our observations were consistent with observations by Li et al. that adopting the ERS/ATS-2022 criteria led to a significant reduction in the BDR of spirometry for both asthma and COPD patients.^21^

A significant number of patients with obstructive lung disease classified as nonresponsive based on spirometry may have a significant BDR in the RV.^22^ The BDR of the RV in our study was similar to that reported by McCartney et al.^22^ Unlike them, we observed that the severity of airflow obstruction and underlying diseases did not influence the presence of BDR in the RV.

The ERS technical standards are silent on whether the presence of the BDR of any of the oscillometry parameters will be considered as BDR. The thresholds for BDR in oscillometry parameters were inconsistent across previous studies and differed from the ERS standard.^23–25^ Park et al. reported that a cutoff lower than the ERS standard had very high sensitivity and specificity for diagnosing BDR of oscillometr y.^14^ Cottee et al. reported that the BDR of oscillometry parameters in asthma patients was twice that of spirometry.^24^ Park et al. reported that the BDR of oscillometry parameters in asthma patients was not different from that in COPD patients.^14^ Unlike ours, Lu et al. observed that the AX of COPD patients had the highest BDR among all oscillometry parameters.^25^ The BDR of X5 and R5 in their COPD patients was much lower than ours. The difference between our and earlier studies could be due to different cutoffs in defining the BDR. The concordance between the BDR of oscillometry parameters and spirometry parameters was poor across all studies, as the two tests measure different respiratory system mechanics. The criteria and prevalence of BDR of Rrs at a higher frequency, i.e., R19, have never been reported. We observed a >40% decrease in R19 in only 1.3% of the cohort.

The PR in spirometry due to bronchodilator-associated bronchoconstriction is well recognized. The PR in spirometry of our study was a little lower than that reported by the COPDGene cohort.^26^ The mechanism of PR in oscillometry is different. Bronchodilators can increase Rrs and decrease Xrs because bronchodilators affect airway caliber and closure.^9^ The criteria for PR in oscillometry is yet to be defined. The prevalence of PR in oscillometry parameters has not been reported earlier. We observed that COPD patients are likely to have more PR in oscillometry parameters. This might be because the FVC of COPD patients usually has a relatively high BDR. To the best of our knowledge, our study is the first to document PR in spirometry and oscillometry in patients with obstructive airway disease. The agreement of BDR of oscillometry and other lung function parameters is weak.

Our study had limitations. The study design limited the functional assessment of the patients. The data were collected from a single center. Because our study was hospital-based, many COPD patients had severe airflow obstruction. We did not distinguish patients with asthma COPD overlapped from COPD patients.

In conclusion, oscillometry parameters worsened with the progression of airflow obstruction, regardless of the underlying disease. The FOT did not differ between asthma and COPD patients with similar airflow obstructions, except for R5 and R19. The distribution of high oscillometry parameters between asthma and COPD patients with identical airflow obstructions was not different. The severity of abnormalities in oscillometry parameters is not affected by the underlying disease but rather by the severity of airflow obstruction. Asthma patients also experience expiratory flow limitations at tidal breaths regardless of the severity of airflow obstruction. Oscillometry parameters of both asthma and COPD patients had a significantly lower BDR than all spirometry parameters, including MMEF.

## Data Availability

All data produced in the present study are available upon reasonable request to the authors

## Author Contributions

Conceived, interpretation of results, drafting the manuscript by SD. Data collection by AS. Both authors approved the manuscript and decided to submit it for publication.

## Abbreviations list

ATS: American Thoracic Society
BDR: Bronchodilator responsiveness
COPD: Chronic obstructive pulmonary disease
EFL_T_: Expiratory flow limitation at tidal breaths
ERS: European Respiratory Society
FEV_1_: Forced expiratory volume 1 s
FVC: Forced vital capacity
MMEF: Maximal mid-expiratory flow
PR: Paradoxical responsiveness
R5: Respiratory system resistance at 5 Hz
R5-19: Difference in respiratory resistance between 5 Hz and 19 Hz
Rrs: Respiratory system resistance
sRaw: Specific airway resistance
X5: respiratory system reactance at 5 Hz
Xrs: Respiratory system reactance
ΔX5: Difference between inspiratory and expiratory reactance at 5 Hz

